# Intermittent Hypoxemia and Brain Injury Biomarker S100B in Preterm Infants

**DOI:** 10.1101/2025.09.05.25334486

**Authors:** Elie G. Abu Jawdeh, Linda J. Van Eldik, Jennifer Stevenson, Abhijit Patwardhan, Philip M. Westgate, Richard Martin, Henrietta S. Bada

## Abstract

**Background:** Intermittent hypoxemia (IH) is common in preterm infants and linked to brain injury. S100B is a glial-derived protein that rises early after neural injury and can be measured noninvasively in urine. We evaluated the relationship between IH burden and urinary S100B in preterm infants ≤32 weeks’ gestation.

**Methods:** Preterm infants ≤32 weeks’ gestation were prospectively enrolled. Oxygen saturation was continuously monitored, and IH profiles were quantified using validated algorithms. Urine S100B was measured by ultrasensitive immunoassay. Infants with severe intraventricular hemorrhage were excluded. Spearman correlations examined associations between IH metrics and urinary S100B, overall and by gestational age subgroups.

**Results:** Twenty-one infants contributed 53 urine samples. Higher urinary S100B correlated with greater IH frequency, percent time in hypoxemia, longer event duration, and lower nadir saturations (all p <0.05). Short events showed the strongest correlations for frequency (*ρ* = 0.50) and percent time (*ρ* = 0.54), while longer events correlated most strongly with nadir (*ρ* = –0.66). Extremely preterm infants demonstrated stronger associations for nadir and duration; very preterm infants only for percent time. S100B increased stepwise across IH burden tertiles.

**Conclusions:** Urinary S100B increases with IH burden, with patterns varying by gestational age and event duration. Urinary S100B may provide an early, noninvasive biomarker of IH-related brain injury in preterm infants.

## Background

Infants born at ≤32 weeks’ gestation experience the highest burden of intermittent hypoxemia (IH) due to immature respiratory control and chronic lung disease^1^. IH, characterized by frequent, episodic drops in oxygen saturation, can occur hundreds of times per day in this population^1–3^. Over the past decade, evidence from both preterm infants and neonatal animal models has consistently linked IH to neurological injury, including white matter injury, neurodevelopmental impairment (NDI), retinopathy of prematurity (ROP), and heightened inflammation^3–7^. However, brain injury in these infants is often not recognized until months or years later during follow-up visits, when the damage may already be irreversible. Evidence from adults with obstructive sleep apnea (OSA), a disorder defined by recurrent IH, provides further support that IH contributes to brain injury as well. In OSA, repetitive hypoxic episodes are linked to both structural and functional brain injury and to elevated levels of systemic brain injury biomarkers^8–14^. In preterm infants, IH exposure is more frequent, more prolonged, and occurs during a critical period of brain development, yet no early biomarkers are available in clinical care to identify IH-related brain injury. At present, injury is examined only months to years later through neurodevelopmental assessments^3,4,6^, a delay that hinders both prognosis and the evaluation of IH-targeted therapies. Identifying a reliable systemic biomarker of IH-related brain injury could allow early risk stratification, timely intervention, and serve as a surrogate endpoint for interventional studies.

S100B is a glial-derived calcium-binding protein expressed predominantly by astrocytes, with measurable concentrations in blood and urine^15–17^. In neonates, elevated S100B during the early postnatal period has been associated with adverse outcomes including periventricular leukomalacia (PVL) ^15,18–23^. In adults with OSA, multiple studies have shown that systemic brain injury biomarkers, including S100B, are elevated compared with controls and correlate with disease severity^24–26^. Establishing whether S100B reflects IH-related brain injury in preterm infants could close a critical gap by providing a feasible, early, and noninvasive biomarker for both clinical and research applications. In this study, we assessed for the first time in preterm infants the relationship between IH burden and S100B levels.

## Methods

### Study Population

We prospectively enrolled preterm infants born at ≤32 weeks’ gestational age (GA) and admitted to the neonatal intensive care unit (NICU) at the University of Kentucky. Infants with severe intraventricular hemorrhage (grade III or IV) were excluded. The study was approved by the University of Kentucky Institutional Review Board, and written informed consent was obtained from parents prior to enrollment.

### Intermittent Hypoxemia Monitoring and IH Profile

Infants were continuously monitored using a research-grade pulse oximeter (Rad-7, Masimo Corp., Irvine, CA) with a 1 Hz sampling rate and 2-second averaging time to enhance resolution. Research alarms were silenced to reduce alarm fatigue for nursing staff. IH profiles were calculated using custom software developed by our group^27^. This validated automated analysis algorithm quantifies IH profile with excellent agreement with manual annotation (r = 0.99)^27^. The IH profile for this study includes frequency, percent time in hypoxemia, duration of events, nadir (severity), mean SpO₂, and categorization of events by <1 min and ≥1 min duration. The IH profile was calculated using thresholds of SpO₂ <80% and <85% for the 24-hour period prior to biomarker measurement. Infants without contemporaneous physiologic data in this 24-hour window were excluded.

### Urine Sample Collection and S100B Analysis

Urine samples were collected at multiple time points during the NICU stay depending on gestational age. A cotton ball was placed in the diaper and subsequently transferred to a sterile container by bedside nursing staff. Samples were processed promptly, aliquoted, and stored at – 80 °C until batched analysis at the University of Kentucky Center for Clinical and Translational Science Biomarker Core Facility. S100B concentrations were measured using an ultrasensitive electrochemiluminescence immunoassay (Meso Scale Discovery, Rockville, MD) optimized for low sample volumes. Urinary S100B values were not corrected for creatinine clearance, given prior evidence that urinary levels are not affected by renal function^28^.

### Clinical Data Collection

Baseline demographic and clinical characteristics were abstracted from medical records.

### Statistical Analysis

Baseline demographic and clinical characteristics are presented as medians (interquartile ranges) or counts (percentages). Correlations between IH parameters and urinary S100B were examined using Spearman correlation coefficients, with analyses weighted by the number of urine samples contributed per infant. Analyses were performed for the overall cohort and separately for extremely preterm (<28 weeks GA) and very preterm (28–31 weeks GA) infants. In the overall cohort, correlations were also calculated separately for short (<1 min) and long (≥1 min) desaturation events. Additionally, samples were categorized into tertiles based on IH burden. Differences in S100B concentrations across tertiles were assessed using the Kruskal–Wallis test, with post-hoc pairwise comparisons by the Mann–Whitney U test. A two-tailed p-value <0.05 was considered statistically significant. Analyses were performed using SPSS [version 30, IBM Corp., Armonk, NY].

## Results

### Study Cohort

A total of 29 preterm infants born at ≤32 weeks’ GA were enrolled; after exclusions (4 infants grade III-IV IVH, 4 infants without IH data at the time of urine samples collection), 21 infants contributed 53 urine samples. The median GA was 28.4 weeks and the median birth weight was 1,060 g. Baseline demographic and clinical characteristics are shown in Table 1.

**Table 1.** Demographics and Baseline Characteristics.

| **Table 1. Demographics and Baseline Characteristics** |  |
| --- | --- |
| Gestational Age (weeks) | 28.4 [26.3–32.4] |
| Birth weight (grams) | 1060.0 [855.0–1300.0] |
| Samples per patient | 2.0 [1.0–4.0] |
| Postnatal age at sample collection (days) | 36.33 [23.0–46.4] |
| Male | 12 (57.1%) |
| Non-Hispanic White | 19 (90.4%) |
| Cesarean Section | 17 (81.0%) |
| Multiple Gestation | 8 (38.1%) |
| Prenatal Steroids | 20 (95.2%) |
| Respiratory Distress Syndrome | 18 (85.7%) |
| Intraventricular Hemorrhage (IVH) |  |
| No IVH | 16 (76.2%) |
| Grade 1 IVH | 4 (19.0%) |
| Grade 2 IVH | 1 (4.8%) |
| Median [interquartile range], number (percent) |  |

### IH Burden and Urinary S100B

In the full cohort (Table 2), greater percent time with SpO₂ <85% and SpO₂ <80%, higher IH frequency at both thresholds, longer IH duration, and lower nadir SpO₂ were all associated with higher urinary S100B concentrations (all p <0.05). The strongest correlations were observed for nadir SpO₂ (*ρ* = –0.56 at SpO₂ <85%; *ρ* = –0.62 at SpO₂ <80%) and for IH frequency (*ρ* = 0.55 at both thresholds, p <0.001). Mean SpO₂ was inversely correlated with S100B (*ρ* = –0.38, 95% CI –0.60 to –0.11, p = 0.005).

**Table 2.** Correlations between urinary S100B and the IH profile.

| **Table 2. Correlations between urinary S100B and the IH profile** |  |  |
| --- | --- | --- |
| **IH Profile Measure** | **Spearman's $\rho$ (95% CI)** | **p-value** |
| % Time — SpO<sub>2</sub> <85% | 0.44 (0.19 to 0.64) | 0.001 |
| % Time — SpO<sub>2</sub> <80% | 0.52 (0.29 to 0.70) | <0.001 |
| IH Frequency — SpO<sub>2</sub> <85% | 0.55 (0.33 to 0.72) | <0.001 |
| IH Frequency — SpO<sub>2</sub> <80% | 0.55 (0.32 to 0.72) | <0.001 |
| IH Nadir — SpO<sub>2</sub> <85% | −0.56 (−0.72 to −0.33) | <0.001 |
| IH Nadir — SpO<sub>2</sub> <80% | −0.62 (−0.76 to −0.41) | <0.001 |
| IH Duration — SpO<sub>2</sub> <85% | 0.28 (0.01 to 0.52) | 0.040 |
| IH Duration — SpO<sub>2</sub> <80% | 0.54 (0.31 to 0.71) | <0.001 |
| Mean SpO<sub>2</sub> | −0.38 (−0.60 to −0.11) | 0.005 |

### Event Duration

When IH events were stratified by duration (Table 3), short events (<1 min) showed the strongest positive correlations with S100B for percent time (*ρ* = 0.54, p <0.001) and frequency (*ρ* = 0.50, p <0.001). Long events (≥1 min) showed the strongest negative association with nadir SpO₂ (*ρ* = – 0.66, 95% CI –0.80 to –0.45, p <0.001).

**Table 3.** Correlations between urinary S100B and the IH profile by event duration.

| **Long events ($\geq 1$ min)** |  |  |
| --- | --- | --- |
| **IH Profile Measure** | **Spearman's $\rho$ (95% CI)** | **p-value** |
| % Time — SpO<sub>2</sub> <85% | 0.31 (0.04 to 0.54) | 0.023 |
| % Time — SpO<sub>2</sub> <80% | 0.35 (0.08 to 0.57) | 0.011 |
| IH Frequency — SpO<sub>2</sub> <85% | 0.28 (0.00 to 0.52) | 0.043 |
| IH Frequency — SpO<sub>2</sub> <80% | 0.34 (0.07 to 0.56) | 0.013 |
| IH Nadir — SpO<sub>2</sub> <85% | −0.66 (−0.80 to −0.45) | <0.001 |
| IH Nadir — SpO<sub>2</sub> <80% | −0.64 (−0.79 to −0.40) | <0.001 |
| **Short events (<1 min)** |  |  |
| **IH Profile Measure** | **Spearman's $\rho$ (95% CI)** | **p-value** |
| % Time — SpO<sub>2</sub> <85% | 0.54 (0.31 to 0.71) | <0.001 |
| % Time — SpO<sub>2</sub> <80% | 0.54 (0.31 to 0.71) | <0.001 |
| IH Frequency — SpO<sub>2</sub> <85% | 0.50 (0.26 to 0.69) | <0.001 |
| IH Frequency — SpO<sub>2</sub> <80% | 0.52 (0.28 to 0.70) | <0.001 |
| IH Nadir — SpO<sub>2</sub> <85% | −0.28 (−0.52 to −0.00) | 0.041 |
| IH Nadir — SpO<sub>2</sub> <80% | −0.41 (−0.62 to −0.15) | 0.002 |

### Gestational Age Stratification

When stratified by GA (Table 4), extremely preterm infants (<28 weeks) demonstrated significant correlations between S100B and nadir SpO₂ (*ρ* = –0.46 at SpO₂ <85%; *ρ* = –0.46 at SpO₂ <80%, both p <0.01) as well as IH duration (*ρ* = 0.37–0.42, p <0.05). In very preterm infants (28–31 weeks), significant associations were observed for percent time with SpO₂ <85% (*ρ* = 0.45, p = 0.038) and SpO₂ <80% (*ρ* = 0.45, p = 0.041), whereas correlations with nadir SpO₂ and IH duration were not significant.

**Table 4.** Correlations between urinary S100B and the IH profile by gestational age group.

|  |  |  |  |  |
| --- | --- | --- | --- | --- |
| **IH Profile Measure** | **Extremely preterm**<br>**ρ (95% CI)** | **p-value** | **Very preterm**<br>**ρ (95% CI)** | **p-value** |
| **% Time — SpO<sub>2</sub> <85%** | 0.21 (−0.16 to 0.53) | 0.243 | 0.45 (0.02 to 0.75) | 0.038 |
| **% Time — SpO<sub>2</sub> <80%** | 0.51 (0.18 to 0.73) | 0.003 | 0.45 (0.01 to 0.74) | 0.041 |
| **IH Frequency — SpO<sub>2</sub> <85%** | 0.37 (0.01 to 0.64) | 0.037 | 0.33 (−0.13 to 0.68) | 0.139 |
| **IH Frequency — SpO<sub>2</sub> <80%** | 0.40 (0.05 to 0.66) | 0.022 | 0.29 (−0.18 to 0.65) | 0.201 |
| **IH Nadir — SpO<sub>2</sub> <85%** | −0.56 (−0.76 to −0.25) | 0.001 | −0.20 (−0.59 to 0.26) | 0.376 |
| **IH Nadir — SpO<sub>2</sub> <80%** | −0.62 (−0.80 to −0.33) | <0.001 | −0.36 (−0.69 to 0.10) | 0.106 |
| **IH Duration — SpO<sub>2</sub> <85%** | 0.56 (0.25 to 0.76) | 0.001 | −0.07 (−0.50 to 0.39) | 0.767 |
| **IH Duration — SpO<sub>2</sub> <80%** | 0.80 (0.62 to 0.90) | <0.001 | 0.29 (−0.18 to 0.65) | 0.201 |
| **Mean SpO<sub>2</sub>** | −0.24 (−0.55 to 0.13) | 0.187 | −0.38 (−0.70 to 0.08) | 0.090 |

### Tertile Analyses of IH Burden

When samples were grouped into tertiles by IH burden (Figure 1, n=7 infants per tertile), urinary S100B concentrations increased stepwise across tertiles (p <0.0001). Significant differences were observed between tertile 1 and tertile 3 (p <0.0001) and between tertile 2 and tertile 3 (p <0.05), consistent with a dose–response relationship between IH burden and S100B release.

**Figure 1.**
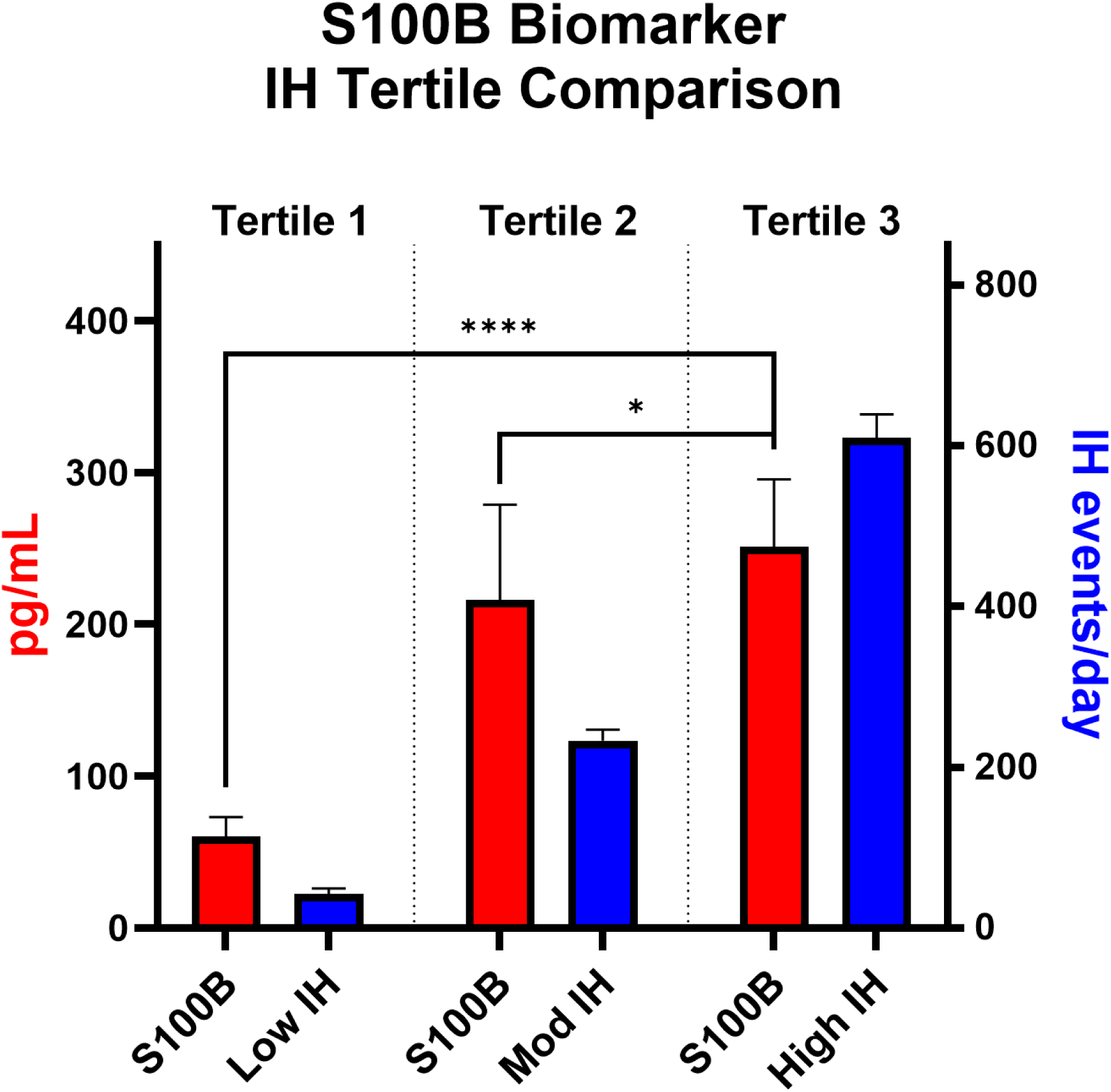
Increase in S100B with higher IH burden. Samples were divided into 3 tertiles based on IH frequency: low, moderate (mod), and high IH. Urinary S100B (red bars) levels increased stepwise across tertiles, in parallel with increasing IH frequency (blue bars). Significant differences in S100B levels were observed across tertiles (Kruskal–Wallis, p <0.0001). ****p <0.0001, *p <0.05 (Mann–Whitney). Differences across IH groups (blue bars) were all p <0.001 (asterisks not shown to avoid crowding). Data shown as mean ± SEM.

## Discussion

In this study, we demonstrate for the first time that IH burden in preterm infants is associated with elevations in urinary S100B, a biomarker of brain injury. These findings provide early evidence that S100B may serve as a feasible, early, and noninvasive marker of IH-related brain injury, addressing a critical gap in neonatal care.

The relationship between IH and brain injury is supported by mounting evidence from both neonatal and adult literature. Neonatal animal models show IH induces brain injury through white matter injury, impairs myelination, and increases inflammation^29^. Prior clinical studies have also linked IH to adverse outcomes in preterm infants. For example, a post-hoc analysis of 1,019 extremely preterm infants from the Canadian Oxygen Trial showed that those in the highest decile of IH exposure had a significantly higher risk of late death or impairment at 18 months compared with those in the lowest decile (56% vs. 36.9%; relative risk [RR] 1.53, 95% CI 1.21–1.94)^4^. Notably, this association was driven by longer IH episodes (>1 minute; RR 1.66), whereas shorter episodes (<1 minute) did not confer added risk (RR 1.01)^4^. Kim et al. in a single center study showed that each additional hour of total IH with SpO₂ ≤80% was associated with a 2.7% increase in the odds of severe NDI at 18–24 months corrected age^6^. Similarly, in a cohort of 210 infants <31 weeks’ gestation, cumulative IH of longer duration during the second month of life was associated with a twofold increased risk of retinopathy of prematurity (ROP) (adjusted odds ratio 2.01 per 3.8-second increase in IH)^5^. Together, these data reinforce that recurrent IH contributes directly to preterm brain injury and subsequent NDI and provide strong rationale for biomarker-based approaches such as urinary S100B to capture injury risk.

S100B is released rapidly after brain injury, and can be detected systemically in both blood and urine. In neonates, elevated S100B during the early postnatal period has been associated with brain injury such as periventricular leukomalacia^21,22^. In infants with hypoxic-ischemic encephalopathy (HIE), S100B levels are higher in moderate-to-severe compared with mild HIE and correlate with the severity of encephalopathy, extent of MRI injury, and adverse outcomes including cerebral palsy and death^30–33^.

In adults, S100B is similarly responsive to hypoxia-related exposures. Patients with OSA have elevated serum S100B levels compared with controls, with values correlating to disease severity. Other hypoxic exposures provide additional insights into the relationship between hypoxia and brain injury; including study of Canadian Armed Forces aviators and competitive breath-hold divers, who experience hypoxemia exposure, exhibit elevated levels of multiple brain injury biomarkers^34,35^. Another study in recreational scuba divers measured serum S100B before and after three consecutive dives, showing mean levels rising from 0.06 μg/L at baseline to 0.086 μg/L post-dive (p = 0.057)^36^. Taken together, these studies indicate that brief hypoxic exposures can provoke measurable elevations in brain injury biomarkers. Hence, we speculate it is biologically plausible that frequent and persistent IH episodes experienced by preterm infants during a vulnerable period of brain development drive measurable brain injury, as captured here by urinary S100B.

Our results extend this finding by assessing the relationship between IH and S100B in preterm infants. Notably, we observed that increased IH burden correlated with elevations in S100B in a stepwise manner across tertiles, suggesting a dose–response relationship. Although overall longer IH duration correlated with higher S100B, the relative contribution of short versus long events varied by metric: short events showed the strongest associations for frequency, whereas long events showed the strongest associations with nadir severity. These findings suggest that both brief, frequent IH events and prolonged events with deeper nadirs may be injurious. This differs from Poets et al., who reported significance only for events ≥1 minute^4^. The discrepancy may reflect methodological differences, as their study used monitors with a long averaging time (16 seconds), which may overestimate longer IH events while merging brief events^3,27,37^. In contrast, our use of a 2-second averaging time enabled high-resolution quantification of both short and longer IH events^3,27,37^. Correlations between IH and S100B were also more prominent in extremely preterm compared with very preterm infants, highlighting potentially increased brain susceptibility at earlier gestational ages. Furthermore, longer IH episodes contributed more strongly to S100B elevations than shorter episodes, mirroring prior outcome studies where prolonged events, rather than brief dips, were most predictive of later impairment. These findings support that urinary S100B may be sensitive to the frequency, severity, and duration of IH exposure, making it a promising biomarker for mechanistic and interventional studies.

If validated in larger studies, urinary S100B could transform how IH-related brain injury is detected and monitored. S100B concentrations are not influenced by renal function^28^, hence, its noninvasive nature and feasibility for serial sampling make it ideally suited for the neonatal intensive care setting, where repeated blood sampling is limited in this vulnerable population. A clinically available biomarker could enable early risk stratification, guide neuroprotective interventions, and serve as a surrogate endpoint for IH-targeted therapies.

Several limitations warrant consideration. First, this was an exploratory analysis in a modest sample size. Second, S100B can be elevated in other conditions affecting the brain; however, the major confounder such as severe intraventricular hemorrhage was excluded. Third, our sample size limited adjustment for multiple confounders. Instead, we stratified by gestational age as the major factor associated with developmental differences. In addition, this study did not evaluate whether urinary S100B elevations translate into later neurological outcomes in this setting, which is essential to establish predictive validity in the future. Finally, we focused on urinary rather than blood S100B measurements for pragmatic reasons, given limited blood volumes in this population. Urine-based assays are practical in this setting, as S100B is not affected by renal function and can be more feasibly implemented as a biomarker in clinical practice.

In summary, this is the first study to demonstrate that urinary S100B increases with greater IH burden in preterm infants. Ongoing studies will be essential to validate these findings and establish clinically meaningful thresholds for S100B. Ultimately, urinary S100B holds promise as an early, noninvasive biomarker of IH-related brain injury in preterm infants, and its incorporation into neonatal care may enable earlier detection, risk stratification, and targeted intervention to mitigate brain injury.

## Data Availability

All data produced in the present study are available upon reasonable request to the corresponding author.

## Acknowledgements

We are deeply grateful to the families who participated in this study. We extend our special thanks to the research nurse coordinators for their dedication and invaluable contributions.

## Author Contributions

E.G.A., L.J.V.E., and H.S.B. conceived the study. E.G.A. conducted and supervised enrollment. E.G.A. and A.P. supervised bedside data collection and processing. J.S. performed biomarker analyses. E.G.A., L.J.V.E., J.S., R.J.M., P.M.W., A.P., and H.S.B. interpreted the data. P.M.W. supervised statistical analyses, and E.G.A. performed the statistical analysis. E.G.A., J.S., and H.S.B. provided resources. E.G.A. drafted the manuscript. All authors reviewed and edited the manuscript and gave final approval of this version to be published.

## Consent Statement

Under a protocol approved by the Institutional Review Board of the University of Kentucky, informed consent was obtained by the research team from each subject’s parent prior to enrollment.

## Data Availability Statement

The data that support the findings of this study are available from the corresponding author upon reasonable request.

